# The Influence of Contextual Factors on the Initial Phases of the COVID-19 Outbreak across U.S. Counties

**DOI:** 10.1101/2020.05.13.20101030

**Authors:** Wolfgang Messner, Sarah E. Payson

## Abstract

**Objectives:** To examine the influence of county- and state-level characteristics on the initial phases of the COVID-19 outbreak across U.S. counties up to April 14, 2020.

**Methods:** We used a statistical exponential growth model for the outbreak. Contextual factors at county- and state-level were simultaneously tested with a multilevel linear model. All data was publicly available.

**Results:** Collectivism was positively associated with the outbreak rate. The racial and ethnic composition of counties contributed to outbreak differences, affecting Black/African and Asian Americans most. Counties with a higher median age had a stronger outbreak, as did counties with more people below the age of 18. Higher income, education, and personal health were generally associated with a lower outbreak. Obesity was negatively related to the outbreak. Smoking was negatively related, but only directionally informative. Air pollution was another significant contributor to the outbreak, but population density did not give statistical significance.

**Conclusions:** Because of high intrastate and intercounty variation in contextual factors, policy makers need to target pandemic responses to the smallest subdivision possible, so that countermeasures can be implemented effectively.

## Introduction

First reports of a pneumonia of unknown etiology emerged in Wuhan, China, on December 31, 2019. The extremely contagious virus was identified as severe acute respiratory syndrome coronavirus 2 (SARS-CoV-2), and spread quickly beyond Wuhan. In the U.S., the first case of COVID-19, the disease caused by SARS-CoV-2, was reported on January 22, 2020. Despite unprecedented government action, the number of cases in the U.S. grew to over 593000 as of April 14, and crossed one million on April 28.^1^

The local press and epidemiological research alike have reported local differences in the outbreak.^2^ A community’s susceptibility to any virus is determined by a variety of factors, inter alia, biological determinants, demographic profiles, and socioeconomic characteristics.^3^ These factors vary significantly across the U.S.; for instance, COVID-19 fatalities in New York, an epicenter of the outbreak in the U.S., disproportionally affect males and people belonging to older age groups, from Black/African and Hispanic ethnicities, and with certain comorbidities.^4^

As of May 09, 2020, more than half of COVID-19 data reported by the Centers for Disease Control and Prevention (CDC) are missing race and ethnicity disaggregation; other individual variables are lacking as well. To understand local differences in the outbreak and risk of contracting COVID-19, we therefore deploy an ecological analysis using contextual factors. A two-level hierarchical linear model with full maximum likelihood allows us to simultaneously test and disentangle county- and state-level effects.

Our study contributes to various strands of current COVID-19 research. First, we note that contextual factors influence the COVID-19 outbreak. Because significant variations in the outbreak exist between states and counties within a state (see Figures 1 and 2),^2^ we recommend policy makers to look at pandemics from the smallest subdivision possible for effective implementation of countermeasures and provision of critical resources. Second, we develop an understanding of how regional cultural differences relate to outbreak variations, driven by specific psychological functioning of individuals and the enduring effects of such differences on political processes, governmental institutions, and public policies.^5,6^ Third, we debunk rumors that a state’s leadership, as expressed by the political party in control or the gender of its governor, has a statistically significant influence on the outbreak.^7^ Fourth, we identify how the virus affects counties differently, depending on their demographic profile. Fifth, while good personal health is generally associated with a lower risk, we identify the prevalence of obesity and smoking in counties to be negatively related with the outbreak. Sixth, while previous studies link air pollution to the death rate, we show that it also contributes to the number of cases.

**Figure 1:**
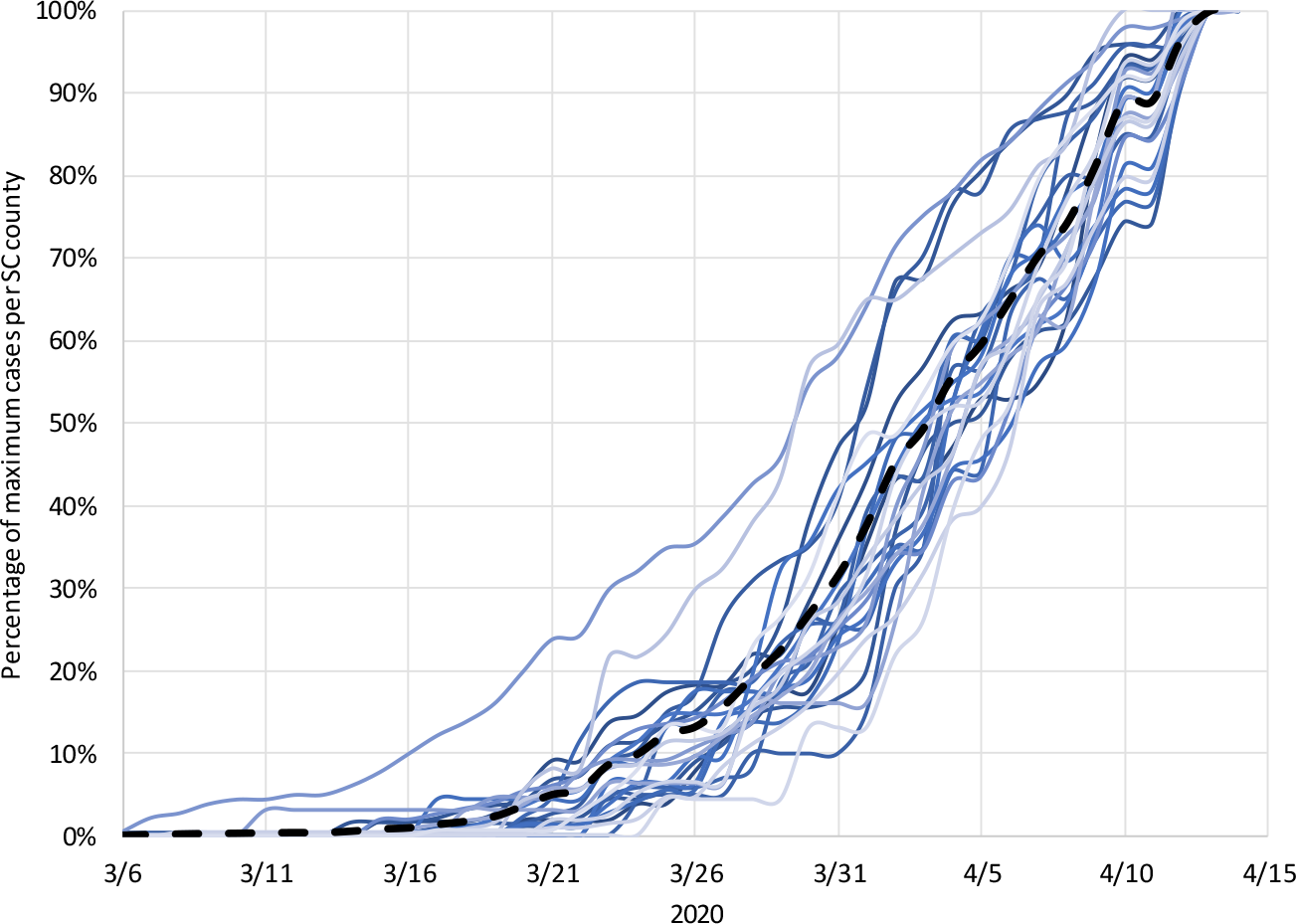
Epidemic days at county level (South Carolina) The spaghetti lines trace the COVID-19 outbreak in South Carolina (black dashed line) and the counties (straight lines) as a percentage of the cases reported on April 14, 2020. Cases unallocated to a county due to lack of information are included in the state line; counties with less than 20 reported cases are not shown in the diagram.

**Figure 2:**
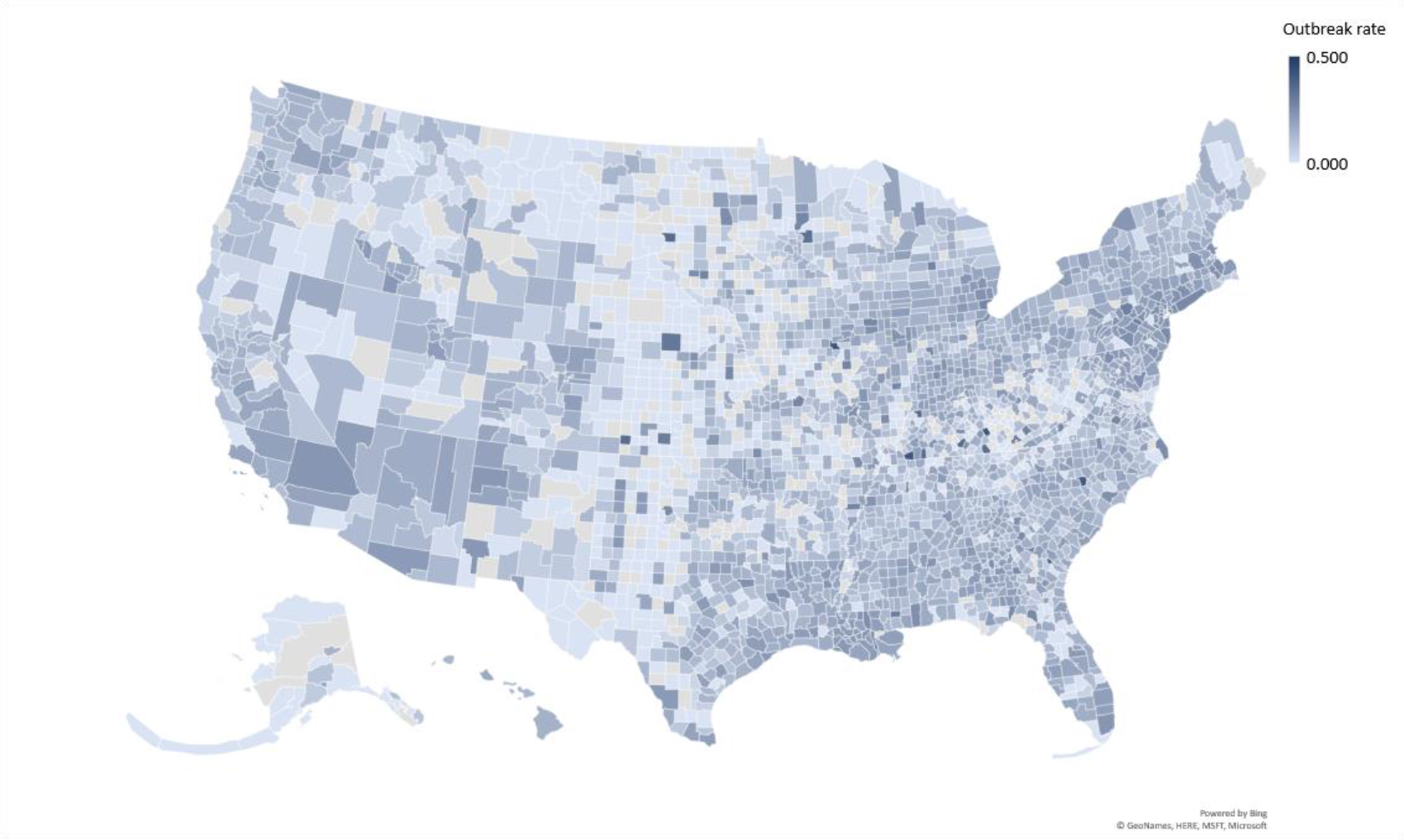
Variation in outbreak rates at U.S. county level. This geo map reveals a large variation in outbreak rates at U.S. county level (April 14, 2020). Lighter colors signify that the pandemic has a slower relative growth rate, and darker colors point to a faster growth.

## Methods

We now explain the estimation of the outbreak rate, and the reasons for including certain contextual factors; Table 1 summarizes the data sources.

**Table 1:** Variables and descriptive statistics.

| Variable | Primary source | Secondary source | N | Year(s) | Median | Minimum; maximum | Std. Dev. |
| --- | --- | --- | --- | --- | --- | --- | --- |
| <u>State institutions</u> |  |  |  |  |  |  |  |
| Party control | WIK |  | 50 | 2020 |  |  |  |
| Gender of governor | WIK |  | 50 | 2020 |  |  |  |
| Government spending | SIP | Census Bureau | 51 | 2015 | 10.059 | 16.553 | 3.186 |
| <u>People cultural values</u> |  |  |  |  |  |  |  |
| Collectivism | VAN |  | 50 | 1997 | 49.500 | [31; 91] | 11.336 |
| <u>Racial composition</u> |  |  |  |  |  |  |  |
| Black & African American | CHR | Census population est. | 3130 | 2018 | 2.251 | [0.512; 85.414] | 14.370 |
| Native American | CHR | Census population est. | 3141 | 2018 | 0.640 | [0.000; 92.515] | 7.600 |
| Asian American | CHR | Census population est. | 3141 | 2018 | 0.736 | [0.000; 43.357] | 2.953 |
| Native Hawaiian | CHR | Census population est. | 3141 | 2018 | 0.063 | [0.000; 48.900] | 1.081 |
| Hispanic American | CHR | Census population est. | 3141 | 2018 | 4.405 | [0.610; 96.360] | 14.273 |
| <u>Income & education</u> |  |  |  |  |  |  |  |
| Household income | CHR | Small area income and poverty est. | 3140 | 2018 | 50547.500 | [15.229; 140382] | 14124.747 |
| Nonproficiency in English | CHR | Census population est. | 3141 | 2014-18 | 0.748 | [0.000; 51.77] | 3.720 |
| Math grade | CHR | Stanford education data archive | 2467 | 2016 | 3.013 | [1.654; 68943] | 3107.118 |
| <u>Other demographics</u> |  |  |  |  |  |  |  |
| Persons under 18 years | CHR | Census population est. | 3140 | 2018 | 22.063 | [7.069; 41.991] | 3.461 |
| Median age | SCP | American community survey | 3142 | 2012-16 | 41.000 | [21.500; 66.000] | 5.355 |
| Female persons | CHR | Census population est. | 3138 | 2018 | 50.301 | [0.192; 76.208] | 2.659 |
| <u>Personal health</u> |  |  |  |  |  |  |  |
| Social associations | CHR | County business patterns | 3141 | 2017 | 11.096 | [0.000; 52.314] | 5.912 |
| Sleep deprivation | CHR | Behavioral risk factor surveillance system | 3141 | 2016 | 32.949 | [8.937; 46.708] | 4.282 |
| Preventable hospitalization | CHR | Mapping Medicare disparities tool | 3098 | 2017 | 4705 | [34; 16851] | 1856.793 |
| Obesity | CHR | United States Diabetes Surveillance System | 3072 | 2016 | 31.300 | [11.800; 47.600] | 4.510 |
| Smoking | CHR | Behavioral Risk Factor Surveillance System | 3072 | 2020 | 17.409 | [6.546; 41.389] | 11.600 |
| <u>External health</u> |  |  |  |  |  |  |  |
| Air pollution | CHR | Environmental public health tracking network | 3107 | 2014 | 9.400 | [2.300; 19.700] | 1.985 |
| Rural area | CHR | Census population est. | 3134 | 2010 | 59.517 | [0.000; 100.000] | 31.437 |
| Food environment | CHR | USDA food environment atlas; Map the meal gap from Feeding America |  | 2015-17 | 7.700 | [0.000; 34.5000] | 1.512 |
| <u>Other confounders</u> |  |  |  |  |  |  |  |
| Density | SCP | American community survey | 3142 | 2012-16 | 44.967 | [0.384; 71615.813] | 1787.612 |
| Temperature <sup>a)</sup> | NCDC |  | 3141 | 2017 | 43.000 | [-14.200; 73.500] | 11.650 |

This table lists the independent variables at both levels of analysis and their provenance.
Notes: CHR: County health rankings, www.countyhealthrankings.org; SCP: Social Capital Project, https://www.jec.senate.gov/public/index.cfm/republicans/2018/4/the-geography-of-social-capital-in-america; NCDC: National Centers for Environment Information, https://www.ncdc.noaa.gov/cag/county/mapping/1/tavg/202003/2/value; VAN: Collectivism index proposed by Vandello & Cohen (1997);<sup>5</sup> WIK: https://en.wikipedia.org/wiki/List_of_United_States_governors.
<sup>a)</sup> The NCDCs do not provide temperature mapping for Hawaii; all Hawaiian counties replaced by historical average data from http://holiday-weather.com/hawaii/averages. All websites accessed in May 2020.

### Outbreak rate

We obtain COVID-19 outbreak data from USA Facts (as of April 14, 2020).^1^ Since January 22, this database has aggregated data from the CDC and other public health agencies. The 21 cases on the Grand Princess cruise ship are not attributed to any counties in California. We discard cases only allocated at the state-level due to lack of information. On average, these are only 308 cases per state, but a few states have as many as 4866 (New Jersey), 1300 (both Rhode Island and Georgia), or 1216 (Washington State). Following approaches by the Institute for Health Metrics and Evaluation at the University of Washington^8^ and the COVID-19 Modeling Consortium at the University of Texas at Austin,^9^ we model the outbreak using the exponential growth equation 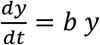, where *b* is a positive constant called the relative growth rate with units of inverse time. Going forward, we simply refer to *b* as the outbreak rate. The shape of the trends in case counts enables us to see differences between counties.^10^ Solutions to this differential equation have the form *y* = *a e^bt^*, where *a* is the initial value of cases *y*. The doubling time *T_d_* can be calculated as 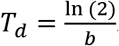. We estimate the outbreak rate for 1987 out of 3142 counties in the 50 U.S. states that have a minimum of 10 reported cases. This is a statistical, but not an epidemiological model, that is, we are neither trying to model infection transmission nor estimate epidemiological parameters, such as the pathogen’s reproductive or attack rate. Instead, we are fitting curves to observed outbreak data at the county level. A change-point analysis using the Fisher discriminant ratio as a kernel function does not show any significant change points in the outbreak, and therefore justifies modeling the COVID-19 outbreak as a phenomenon of unrestricted population growth.^11^ We cannot forecast outbreak dynamics with this statistical approach, though we do not require extrapolated data in our work.

### Cultural values

Culture can be defined as a set of values that are shared in a given social group. While cultural values are often used to distinguish countries,^12^ more than 80% of cultural variation resides within countries.^13^ The original North American colonies were settled by people hailing from various countries, who have spread their influence across mutually exclusive areas. Their distinct cultures are still with us today.^6^ Although today’s U.S. states are not strictly synonymous with these cultural areas, there is abundant evidence that political boundaries can serve as useful proxies for culture.^14^

One of the most useful constructs to emerge from cultural social psychology is the individualism-collectivism bipolarity. It has proven useful in describing cultural variations in behaviors, attitudes, and values. Briefly, individualism is a preference for a loosely knit social framework, whereas collectivism represents a preference for a tightly knit framework, in which its members are interdependent and expected to look after each other in exchange for unquestioning loyalty. While the majority of research on collectivism involves comparing countries,^12^ we use an index developed at state-level solely within the U.S.^5^ Previous studies have shown that the regional prevalence of pathogens and international differences in the COVID-19 outbreak are positively associated with collectivism.^14,15^

### Institutional confounders

In addition to culture, we include various institutional confounders at the state-level, such as the political affiliation of a state’s governor, the gender of the governor, and government spending per capita. Government plays a critical role in policy development and implementation, and so state-level differences could influence the outbreak rate.^16^

### Racial composition

While first systematic reviews about COVID-19 incidences from China relied on ethnically homogenous cohorts,^17,18^ ethnically diverse populations, such as in the U.K. and U.S. may exhibit different susceptibility or response to infection because of socioeconomic, cultural, or lifestyle factors, genetic predisposition and pathophysiological differences. Certain vitamin or mineral deficiencies, differences in insulin resistance or vaccination policies in countries of birth may also be contributing factors.^18^ We include variables measuring the composition of U.S. counties regarding racial and ethnic groups.

### Income and education

Poverty is arguably the greatest risk factor for acquiring and succumbing to disease worldwide, but has historically received less attention from the medical community than genetic or environmental factors. The global HIV crisis brought into sharp relief the vulnerability of financially strapped health systems, and revealed disparities in health outcomes along economic fault lines.^19^ We include the median household income to quantify potential economic disparities between U.S. counties. In addition, we measure non-proficiency in English and math performance of students. Lower educational levels may result in a lower aptitude as it relates to understanding and effectively responding to the pandemic.

### Other demographics

Age and gender also play a potential role in a population’s susceptibility. During the aging process, immune functions decline, rendering the host more vulnerable to certain viruses.^20^ We use the percentage of population below 18 years of age and their median age to determine potential effect of differences in mobility, response, and lifestyle factors. We also control for the percentage of the population that is female, as one COVID-19 study in Italy showed that about 82% of critically ill people admitted into intensive care were men.^21^

### Personal health

Good overall personal health is a general indicator for disease resistance. Additionally, the health belief model suggests that a person’s belief in a personal threat of a disease, together with faith in the effectiveness of behavioral recommendations, predicts the likelihood of the person adopting the recommendation.^22^ We use the percentage of the population that reports insufficient amount of sleep, is obese (as defined by a body mass index above 30), and smokes daily. Given the latter two are publicized risk factors for COVID-19, there is a potential for greater caution following the value-expectancy concepts of the health belief model. Yet, medicinal nicotine has been identified as a potential protective factor against infection by SARS-CoV-2.^23^ We also measure the preventable hospitalization rate (that is, the rate of hospital stays for ambulatory-care sensitive conditions) as a potential indicator of poor personal health and the social association rate (that is, the average number of membership associations), which is generally connected with positive mental health and happiness.

### External health

Previous studies suggest that exposure to pollution can suppress immune responses and proliferate the transmission of infectious diseases,^24^ and that the COVID-19 mortality rate is associated with air pollution.^25^ However, the impact of air pollution on the spread of COVID-19 is not yet known.^24^ We use the 2014 average daily density of fine particulate matter PM_2.5_ to measure air pollution across U.S. counties, and the percentage of population living in rural areas to account for physical distancing being more prevalent in rural areas. In addition, the food environment index reflects access to grocery stores and healthy foods.

### Other confounders

Population density and overcrowding is significant when considering public health crises, facilitating the spread of diseases in developing and developed countries alike.^26^ As the climate is another highly publicized confounder potentially influencing the COVID-19 transmission rate,^27^ we also include each county’s average temperature during February and March 2020. To control for the temporality of the outbreak, we bring in a variable representing the number of days between January 01 and the 10^th^ confirmed case reported.

## Statistical results

To simultaneously test county- and state-level effects of contextual factors on the outbreak rate with cross-level interactions, we estimate a two-level linear model using full maximum likelihood in HLM 7.03 (Figure 3). This accounts for potential similarities in counties within the same state.^24^ We center all predictors around the group mean at level 1 and grand mean at level 2.

**Figure 3.**
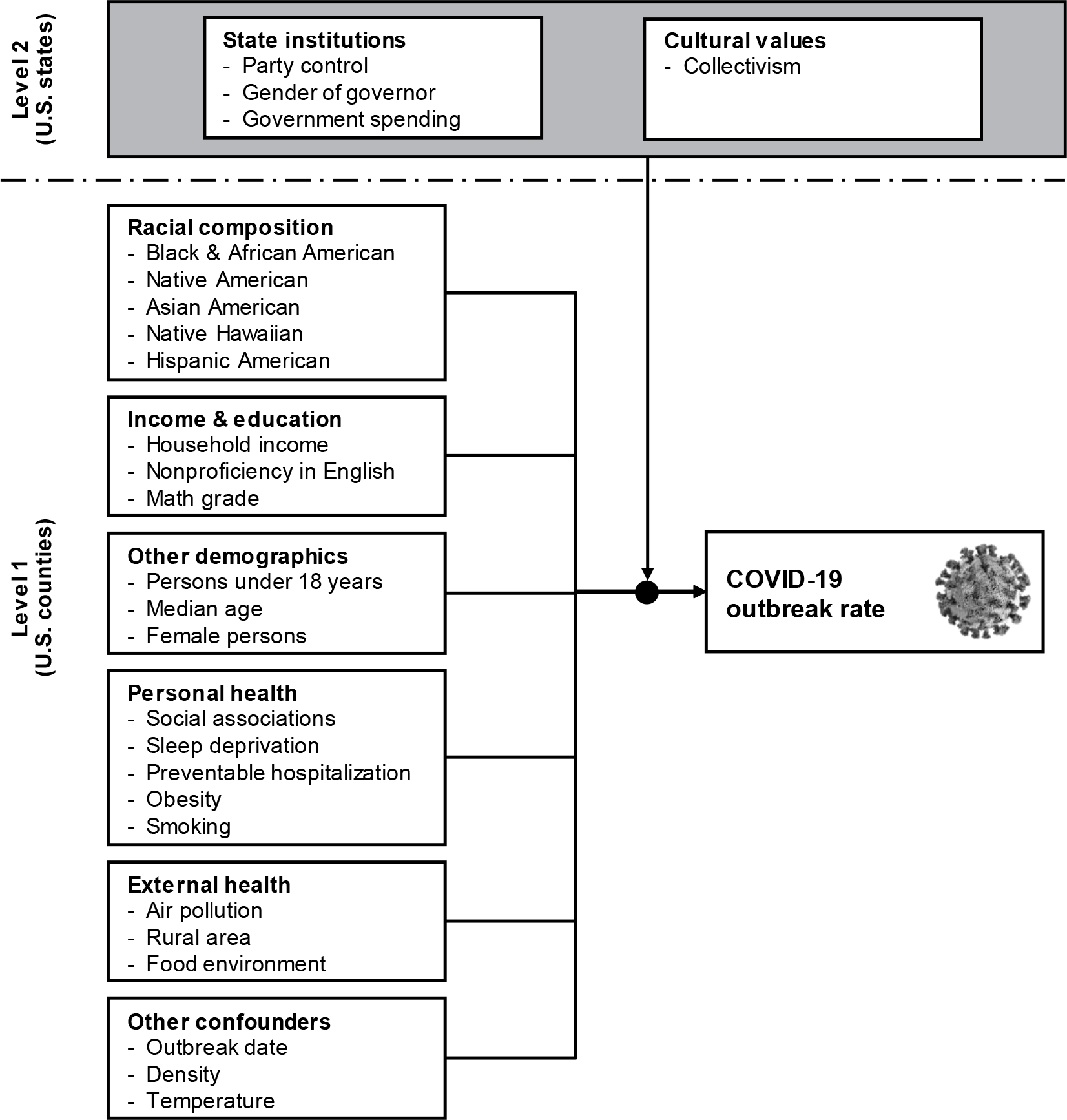
Multilevel research model. This figure details the multi-level research model and the variables used at state- and county-level.

We first estimate a one-way random effects ANOVA (unconditional model), which has an intraclass correlation coefficient (ICC) of 0.243. That is, more than 24% of the variation in the outbreak rate is between states, and about 76% is within the states and between their counties. The variation between states is statistically significant (*u*_0_ = 4.50E-04, *p* < 0.001). We thus deem it prudent to proceed with a multilevel model as follows:

Level 1 (counties): 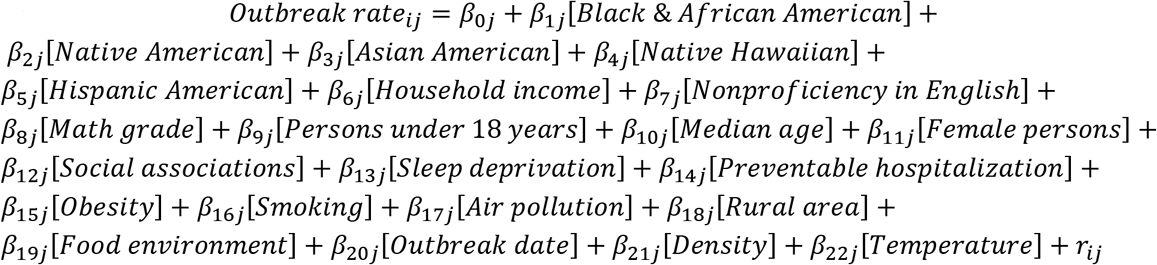
Level 2 (states): 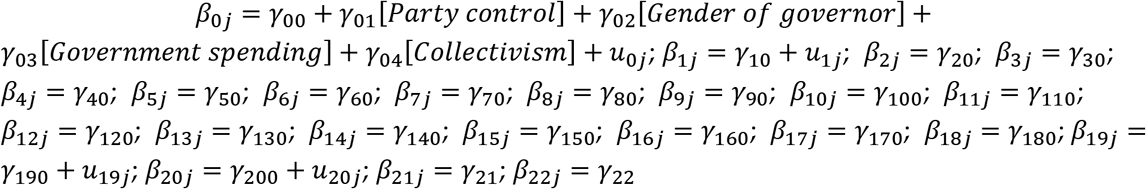

We provide the inter-item correlation matrix in Table 2, and the results of the multilevel model in Table 3. Additionally, we perform several robustness tests to inform our results.

**Table 2:** Inter-item correlation matrix.

|  | b | c | d | e | f | g | h | i | j | k | l | m | n |
| --- | --- | --- | --- | --- | --- | --- | --- | --- | --- | --- | --- | --- | --- |
| a | -0.034 | -0.251 | 0.035 | 0.083 | 0.048 | -0.095 | 0.011 | 0.121 | -0.106 | 0.046 | -0.007 | 0.206 | -0.088 |
| b |  | 0.052 | -0.368 | -0.102 | 0.049 | -0.027 | -0.024 | -0.104 | 0.003 | -0.066 | -0.023 | 0.001 | 0.079 |
| c |  |  | -0.389 | -0.200 | 0.004 | 0.134 | 0.007 | -0.043 | 0.269 | -0.013 | 0.004 | -0.030 | 0.054 |
| d |  |  |  | 0.539 | -0.209 | 0.045 | -0.044 | 0.051 | -0.165 | 0.013 | -0.057 | -0.015 | -0.094 |
| e |  |  |  |  | -0.104 | 0.013 | -0.011 | -0.127 | -0.321 | -0.035 | -0.002 | -0.074 | -0.141 |
| f |  |  |  |  |  | -0.047 | 0.015 | 0.009 | -0.095 | -0.002 | 0.004 | 0.232 | -0.110 |
| g |  |  |  |  |  |  | 0.060 | 0.195 | 0.483 | 0.221 | -0.030 | 0.049 | -0.269 |
| h |  |  |  |  |  |  |  | 0.250 | -0.139 | 0.630 | 0.755 | -0.025 | -0.341 |
| i |  |  |  |  |  |  |  |  | 0.009 | 0.749 | 0.254 | 0.324 | -0.398 |
| j |  |  |  |  |  |  |  |  |  | -0.083 | -0.201 | 0.160 | -0.031 |
| k |  |  |  |  |  |  |  |  |  |  | 0.699 | 0.164 | -0.470 |
| l |  |  |  |  |  |  |  |  |  |  |  | -0.098 | -0.339 |
| m |  |  |  |  |  |  |  |  |  |  |  |  | -0.447 |

|  | o | p | q | r | s | t | u | v | w | x | y | z |
| --- | --- | --- | --- | --- | --- | --- | --- | --- | --- | --- | --- | --- |
| a | -0.006 | -0.083 | -0.002 | -0.007 | 0.115 | 0.028 | 0.057 | 0.025 | -0.156 | 0.014 | -0.095 | 0.258 |
| b | 0.014 | 0.173 | -0.178 | -0.113 | 0.108 | -0.054 | -0.089 | 0.066 | 0.017 | 0.042 | -0.062 | -0.237 |
| c | -0.039 | 0.198 | -0.296 | -0.133 | -0.153 | -0.217 | -0.292 | -0.071 | 0.214 | -0.024 | 0.129 | -0.477 |
| d | 0.128 | -0.272 | 0.557 | 0.301 | 0.203 | 0.138 | 0.407 | -0.030 | -0.241 | -0.116 | 0.045 | 0.715 |
| e | 0.154 | -0.123 | 0.568 | 0.278 | 0.404 | 0.360 | 0.230 | -0.045 | -0.442 | -0.128 | 0.119 | 0.533 |
| f | -0.034 | -0.049 | -0.064 | 0.047 | 0.076 | 0.270 | -0.219 | 0.091 | -0.105 | 0.064 | -0.047 | -0.108 |
| g | 0.058 | -0.169 | -0.046 | -0.137 | -0.362 | -0.315 | 0.097 | -0.501 | 0.087 | -0.448 | 0.553 | 0.033 |
| h | -0.338 | -0.070 | -0.228 | -0.150 | -0.014 | 0.014 | -0.130 | -0.033 | 0.516 | -0.064 | 0.012 | -0.018 |
| i | -0.187 | -0.217 | -0.159 | -0.093 | -0.239 | -0.234 | -0.169 | -0.290 | 0.191 | -0.104 | 0.092 | 0.237 |
| j | 0.064 | -0.051 | -0.327 | -0.322 | -0.518 | -0.666 | -0.031 | -0.384 | 0.309 | -0.353 | 0.246 | -0.276 |
| k | -0.188 | -0.171 | -0.235 | -0.143 | -0.140 | -0.109 | -0.167 | -0.203 | 0.494 | -0.131 | 0.153 | 0.116 |
| l | -0.228 | -0.030 | -0.283 | -0.150 | 0.004 | 0.039 | -0.166 | 0.040 | 0.656 | -0.017 | -0.015 | -0.049 |
| m | 0.138 | -0.117 | -0.020 | 0.103 | 0.144 | 0.004 | 0.055 | -0.190 | -0.063 | -0.028 | -0.062 | 0.021 |
| n | 0.079 | 0.276 | -0.046 | -0.007 | -0.014 | -0.118 | -0.039 | 0.440 | -0.107 | 0.239 | -0.145 | -0.108 |
| o |  | 0.064 | 0.118 | 0.045 | 0.069 | 0.037 | 0.176 | -0.156 | -0.105 | -0.119 | 0.097 | 0.080 |
| p |  |  | -0.259 | -0.031 | 0.041 | -0.076 | -0.092 | 0.142 | 0.063 | 0.206 | -0.065 | -0.288 |
| q |  |  |  | 0.436 | 0.456 | 0.568 | 0.501 | -0.018 | -0.510 | -0.046 | 0.098 | 0.514 |
| r |  |  |  |  | 0.386 | 0.433 | 0.228 | 0.115 | -0.323 | 0.098 | -0.027 | 0.325 |
| s |  |  |  |  |  | 0.611 | 0.289 | 0.283 | -0.296 | 0.246 | -0.228 | 0.213 |
| t |  |  |  |  |  |  | 0.227 | 0.248 | -0.372 | 0.203 | -0.118 | 0.222 |
| u |  |  |  |  |  |  |  | -0.114 | -0.176 | -0.136 | 0.083 | 0.340 |
| v |  |  |  |  |  |  |  |  | 0.001 | 0.518 | -0.369 | -0.064 |
| w |  |  |  |  |  |  |  |  |  | -0.070 | 0.029 | -0.360 |
| x |  |  |  |  |  |  |  |  |  |  | -0.342 | -0.107 |
| y |  |  |  |  |  |  |  |  |  |  |  | 0.043 |
| z |  |  |  |  |  |  |  |  |  |  |  |  |

This table shows the inter-item correlations between the variables at both levels of analysis.
Variables: a: Party control; b: Gender of governor; c: Government spending; d: Collectivism; e: Black & African American; f: Native American; g: Asian American; h: Native Hawaiian; i: Hispanic American; j: Household income; k: Nonproficiency in English; l: Math grade; m: Persons under 18 years; n: Median age; o: Female persons; p: Social associations; q: Sleep deprivation; r: Preventable hospitalization; s: Obesity; t: Smoking; u: Air pollution; v: Rural area; w: Food environment; x: Outbreak date; y: Density; z: Temperature.

**Table 3:** HLM contextual model.

| Fixed effect | Coefficients <sup>a)</sup> | Standard error | Confidence interval | *p* | Effect size <sup>b)</sup> | Reliability | Impact threshold | Confound threshold |
| --- | --- | --- | --- | --- | --- | --- | --- | --- |
| Outbreak rate | 154.947 | 0.003 | [148.685; 161.209] | <0.001 |  | 0.845 |  |  |
| <u>State institutions</u> |  |  |  |  |  |  |  |  |
| Party control <sup>e)</sup> | -2.271 | 0.006 | [-14.382; 9.840] | 0.715 | -4.542 |  | 0.187 | 81.775% |
| Gender of governor <sup>f)</sup> | -3.441 | 0.005 | [-15.034; 8.152] | 0.564 | -8.823 |  | 0.162 | 71.154% |
| Government spending | 0.820 | 0.001 | [-2.318; 3.958] | 0.611 | 0.314 |  | 0.170 | 74.603% |
| <u>People cultural values</u> |  |  |  |  |  |  |  |  |
| Collectivism | 0.998 | 0.000 | [0.351; 1.645] | 0.004 | 0.088 |  | 0.181 | 33.316% |
| <u>Racial composition</u> |  |  |  |  |  |  |  |  |
| Black & African American | 1.158 | 0.000 | [0.725; 1.591] | <0.001 | 0.242 |  | 0.091 | 62.563% |
| Collectivism (cross-level) | -0.049 | 0.000 | [-0.088; -0.010] | 0.014 |  |  | 0.014 | 19.933% |
| Native American | 0.763 | 0.000 | [-0.209; 1.735] | 0.124 | 0.100 |  | 0.011 | 21.581% |
| Asian American | 1.305 | 0.001 | [0.166; 2.444] | 0.025 | 0.441 |  | 0.008 | 12.666% |
| Native Hawaiian | 1.478 | 0.000 | [0.506; 2.450] | 0.538 | 1.369 |  | 0.034 | 68.554% |
| Hispanic American | -0.447 | 0.000 | [-0.915; 0.021] | 0.061 | -0.031 |  | 0.002 | 4.657% |
| <u>Income & education</u> |  |  |  |  |  |  |  |  |
| Household income <sup>c)</sup> | -3.854 | 0.002 | [-7.437; -0.271] | 0.035 | -2.733 |  | 0.004 | 6.957% |
| Nonproficiency in English | 2.090 | 0.001 | [0.547; 3.633] | 0.008 | 0.560 |  | 0.019 | 26.133% |
| Math grade <sup>d)</sup> | -0.002 | 0.000 | [-0.004; 0.000] | <0.001 | 0.000 |  | 0.001 | 1.895% |
| <u>Other demographics</u> |  |  |  |  |  |  |  |  |
| Persons under 18 years | 1.066 | 0.001 | [-0.014; 2.146] | 0.053 | 0.305 |  | 0.001 | 1.375% |
| Median age | 0.657 | 0.000 | [-0.033; 1.347] | 0.062 | 0.107 |  | 0.002 | 4.851% |
| Female persons | 0.167 | 0.001 | [-0.880; 1.214] | 0.755 | 0.054 |  | 0.042 | 84.058% |
| <u>Personal health</u> |  |  |  |  |  |  |  |  |
| Social associations | -2.027 | 0.000 | [-2.911; -1.143] | <0.001 | -0.343 |  | 0.070 | 56.354% |
| Sleep deprivation | 1.557 | 0.001 | [0.412; 2.702] | 0.008 | 0.363 |  | 0.020 | 26.423% |
| Preventable hospitalization | 0.001 | 0.000 | [-0.001; 0.003] | 0.207 | 0.000 |  | 0.024 | 49.022% |
| Obesity | -1.093 | 0.000 | [-1.828; -0.358] | 0.004 | -0.243 |  | 0.027 | 32.698% |
| Smoking | -0.784 | 0.001 | [-3.150; 1.582] | 0.516 | -0.221 |  | 0.033 | 66.888% |
| <u>External health</u> |  |  |  |  |  |  |  |  |
| Air pollution | 3.329 | 0.001 | [1.465; 5.193] | <0.001 | 1.707 |  | 0.043 | 43.962% |
| Rural area | -0.443 | 0.000 | [-0.574; -0.312] | <0.001 | -0.014 |  | 0.127 | 70.332% |
| Food environment | 5.996 | 0.002 | [1.286; 10.706] | 0.016 | 3.945 | 0.284 | 0.015 | 21.384% |
| <u>Other confounders</u> |  |  |  |  |  |  |  |  |
| Outbreak date | 1.912 | 0.000 | [1.322; 2.502] | <0.001 | 0.205 | 0.622 | 0.121 | 69.119% |
| Density | 0.050 | 0.000 | [-0.009; 0.109] | 0.095 | 0.000 |  | 0.007 | 15.037% |
| Temperature | 0.301 | 0.000 | [-0.518; 1.120] | 0.472 | 0.026 |  | 0.031 | 63.291% |

| Random effects | Variance | *df* | $\chi^2$ | *p* |
| --- | --- | --- | --- | --- |
| Variance between state intercepts ($\tau_{00}$) | 4.00E-04 | 43 | 350.334 | <0.001 |
| Variance within states ($\sigma^2$) | 1.26E-03 | | | |

This table provides the detailed results for the multi-level linear model. Run-time deletion reduced the number of level-1 records from 3118 to 1438, and level-2 from 50 to 48.
Notes: <sup>a)</sup> The coefficients are multiplied with 1000 for more intuitive figures. Ditto for the confidence interval. <sup>b)</sup> The effect size is calculated as coefficient / standard deviation, again multiplied with 1000. <sup>c)</sup> The variable for household income is divided by 10000.
<sup>d)</sup> All effects for Math grade are from a separately calculated model because the variable is unavailable for 675 counties across the U.S. Consequently, run-time deletion reduced the number of level-1 records to 1140 and level-2 to 43. This updates some *p*-values, but does not affect the sign of the coefficients. <sup>e)</sup> Party control: 0 = Democratic, 1 = Republican. <sup>f)</sup> Gender of governor: 0 = male, 1 = female.

First, because outbreak rates change over time and their estimation is somewhat sensitive to the starting figure, we alternatively calculate them after 10 and 25 cases in the respective unit, finding a high correlation among the rates. When using the rate after 10 cases, the outbreak date as a control variable changes its sign and loses significance (*p* = 0.065). Notably, the following coefficients gain significance: government spending (*p* = 0.064); temperature (*p* = 0.011). Conversely, the following coefficients lose significance: household income (*p* = 0.989); food environment (*p* = 0.144); density (*p* = 0.709). More importantly, the signs of the coefficients remain the same. The variable outbreak date controls for temporality of the outbreak in the original model (1.912, [1.322; 2.502], *p* < 0.001).

Second, we iteratively include several other contextual variables and logged versions to assess the robustness of the results. But because it is nearly impossible to establish a complete list of confounding variables, we quantify the potential impact of unobserved confounds (Table 3; impact threshold).^28^ For instance, the necessary impact of such a confound for air pollution would be 0.043, that is, to invalidate the variable’s inference on the outbreak rate, a confounding variable would have to be correlated with both the outbreak rate and air pollution at 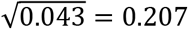. Next, to alleviate concerns that some counties are omitted from the analysis because they are not yet affected by the virus,^24^ we ask how many counties would have to be replaced with unobserved cases for which the null hypothesis is true (i.e., a contextual variables has no influence on the outbreak rate) in order to invalidate the inference.^28^ As Table 3 (confound threshold) shows, 43.962% of the counties would have to be replaced with counties for which the effect is zero in order to invalidate the influence of air pollution. In summary, it can be claimed that the influence of the identified contextual variables on the pandemic is reasonably robust.

Third, a potential omission of relevant variables can lead to multicollinearity issues, which are generally a serious problem in epidemiological studies.^29^ Even though HLM 7.03 checks for multicollinearity, we conduct several additional diagnostics to eliminate any potential issues. In the inter-item correlation matrix (Table 2), the average (absolute) correlation is 0.172, and the highest correlation is 0.754, which is below the typical cutoff of 0.8. Most high correlations exist between racial composition and income and education. Additionally, we conduct a linear regression analysis at level 1 in IBM SPSS 26 (*R*^2^ = 0.495; without variable math grade; pairwise exclusion of missing values), and find that the variable inflation factor (VIF) never exceeds the threshold of 5 (highest being nonproficiency in English, 4.024). The variance-decomposition matrix also does not show any groups of predictors with high values.

Fourth, we rerun our model excluding the 23 counties of the New York metropolitan area. As a COVID-19 hotspot, they could unduly influence our analysis. All signs remain the same, and the following coefficients gain significance: household income (*p* = 0.009); persons under 18 years (*p* = 0.038).

Lastly, we are aware that an accurate estimation and comparison of the outbreak rate across units depends on similar testing strategies, test sensitivities, specificities, and reporting of tests performed vs. individuals tested.^10,30^ Even within the U.S., some states report tests performed and others individuals tested.^30^ The number of tests administered and the number of confirmed cases therefore correlates to varying extents across states.^31^ By using a multi-level model, we aim to accommodate such differences between states.

## Discussion

In the absence of national-level data controlled for location and disaggregated by race and ethnicity, demographics, information about comorbidity and other personal health variables, an ecological analysis provides an alternative way of measuring the disproportionate impact of COVID-19 across the U.S. and among segments of Americans. It may be contrary to expectations that the outbreak rate of a new pathogen, which is able to infect virtually anyone, manifests contextual disparities. But for other conditions, such as HIV and cancer, regional health disparities have been reported before,^32,33^ and with the current study we show that contextual factors in the U.S. also create a variation in COVID-19 cases.

Our analysis indicates that higher outbreak rates can be found in U.S. states characterized by a higher cultural value of collectivism (coefficient 0.998, confidence interval [0.351; 1.645], *p* = 0.004). As Table 2 shows, collectivistic values are more prevalent in counties that are warmer (correlation with temperature 0.715, *p* < 0.001) and have a higher percentage of people with a Black/African background (with Black/African American 0.539, *p* < 0.001). This mirrors findings from international cultural research.^12^ Conversely, we cannot find any statistical evidence that the government spending, the gender of the governor, or the party in control would be in any way linked to the outbreak. This certainly debunks myths spread by the popular media. A disproportionately stronger outbreak of COVID-19 cases can be found in counties with a higher percentage of Black/African (1.158, [0.725; 1.591], *p* < 0.001) and Asian Americans (1.305, [0.166; 2.444], *p* = 0.025), which supports prior infection and mortality studies in the U.S. and U.K.^18,34^ The former counties are also characterized by a higher rate of sleep deprivation (0.568, *p* < 0.001) and warmer temperatures (0.533, *p* < 0.001). The latter have a higher population density (0.553, *p* < 0.001). While we found sleep deprivation to be associated with a higher outbreak rate (1.557; [0.412; 2.702], *p* = 0.008), a positive influence of population density (0.050, [−0.009; 0.109], *p* = 0.095) and temperature (0.301, [−0.518; 1.120], *p* = 0.472) are only directionally informative, but not statistically significant. In the first robustness test, higher average temperatures are positively and significantly related to the outbreak (1.027, [0.235, 1.861], *p* = 0.011), potentially related to more time spent indoor with air conditioning.

Conversely, counties with more Hispanic Americans are less affected by the pandemic, with borderline statistical significance (−0.447, [−0.915; 0.021], *p* = 0.061). We could not find a significant effect for counties with a higher Native American (0.763, [−0.209; 1.735], *p* = 0.124) or Hawaiian population (1.478, [0.506; 2.450], *p* = 0.538) though.

We see that higher income and education levels are associated with a less aggressive outbreak (household income: −3.854, [−7.437; –0.271]; *p* = 0.035; nonproficiency in English: 2.090; [0.547; 3.633]; *p* = 0.008; math grade: –0.002, [−0.004; 000]; *p* < 0.001). In counties with a higher household income, the obesity rate and the percentage of smokers tends to be lower (−0.518, *p* < 0.001 and –0.666, *p* < 0.001 respectively). Both are negatively associated with the outbreak rate. The effect of the obesity rate is highly significant (−1.093, [−1.828; –0.358], *p* = 0.004), but the effect of the percentage of smokers is only directionally informative (−0.784, [−3.150; 1.582], *p* = 0.516). Studies report that people with obesity are at increased risk of developing severe COVID-19 symptoms,^35^ but, to the best of our knowledge, a link to the infection rate has not yet been established. A potential explanation of this is that people with obesity heed the warnings issued by the CDC, and are extra careful in avoiding social contact, in line with the value expectancy concepts of the health belief model.^22^ Other studies report that smoking or medicinal nicotine might be a protective factor against infection by SARS-CoV-2;^23^ our ecological data does not contradict this finding. Many other variables related to good personal health are associated with a slower outbreak (social associations: –2.027, [−2.911; –1.143], *p* < 0.001; sleep deprivation: 1.557, [0.412; 2.702], *p* = 0.008; preventable hospitalization: 0.001, [-0.001; 0.003], *p* = 0.207).

Regarding age-related demographics, we confirm early observations that counties with an older population are more affected by the outbreak, with borderline significance (median age: 0.657, [-0.033; 1.347], *p* = 0.062). Notably, the percentage of persons under 18 years is positively associated with the outbreak rate, again with borderline significance (1.066, [-0.014; 2.146], *p* = 0.053). A possible reason is that younger people physically interact more frequently, closer, and longer with their friends, thus contributing to the spread of the virus. Conversely, we find no effect of differences in gender (0.167, [−0.880; 1.214], *p* = 0.755). None of these demographic variables are strongly correlated with any other variable.

Air pollution is a significant contributor to the outbreak (3.329, [1.465; 5.193], *p* < 0.001), and, concurrently, counties with a rural environment experience a slower outbreak (−0.443, [−0.574; −0.312], *p* < 0.001). This calls for studies linking air pollution to the lethality of COVID-19^24,25^ to include the outbreak rate as a potential confounding variable. Contrariwise, a better food environment is associated with a higher outbreak rate (5.996, [1.286; 10.706], *p* = 0.016). While the food environment index is usually associated with a healthier lifestyle, better access to grocery stores and supermarkets in the vicinity also means more interaction with other people, and thus an increased likelihood of transmission.

As a final point, we want to note that we have presented associations between contextual factors and the COVID-19 outbreak which are consistent with the deliberations leading to our research model. However, these associations, even when statistically significant, are not an inference of causality. Establishing causal inference is, of course, critical for our understanding of and fight against COVID-19, but this represents a direction for further research using more detailed data at the level of individual patients.

## Data Availability

No humans participated in this study. The original data sources are referenced in the section Methods and in Table 1.

